# Quality of life in women with a human papillomavirus-positive screening test, cervical neoplasia, or cervical cancer in Latin America

**DOI:** 10.64898/2026.07.06.26357411

**Authors:** Romina A. Tejada, Arianis Tatiana Ramirez, Annabelle Ferrera, Yessy Cabrera, Carolina Terán, Vania Murillo, Lina Trujillo, Yuli Salgado, Juliana Rodriguez, Mateo Barros, Gino Venegas, María del Pilar Vera, Armando Baena, Guillermo Rodriguez, Andrea Beracochea, Eduardo L. Franco, Maribel Almonte, Talía Malagón, the HPVQoL-LAC Group

## Abstract

**Purpose:** Human papillomavirus (HPV) infection and HPV-associated diseases impact health-related quality of life (HRQOL). We aimed to measure HRQOL in women with HPV-positive test results, cervical intraepithelial neoplasia (CIN), or cervical cancer in Latin America.

**Methods:** We enrolled women aged 18 to 75 years from Bolivia, Colombia, Honduras, Peru, and Uruguay. We used the EQ-5D-5L instrument to calculate EQ-5D index scores with country-specific sets of health preferences when available. We calculated medians and interquartile ranges (IQR) of EQ-5D index and conducted an exploratory analysis comparing HRQOL loss by diagnosis and country with a gamma regression.

**Results:** We present results from 1,073 participants. Median age was 43 years (IQR: 34-52). The EQ-5D index scores by diagnosis were as follows: HPV-positive test alone 0.906 (standard deviation [SD]: 0.129), CIN1: 0.891 (SD: 0.136), CIN2: 0.892 (SE: 0.124), CIN3: 0.870 (SD: 0.138), and cervical cancer: 0.737 (SD: 0.268). HRQOL was associated with age, diagnosis, and country; there was a significant decreasing trend in HRQOL with worsening health state. Women with cervical cancer had a 3.19-fold higher HRQOL loss than women with an HPV-positive test alone or CIN1. We also found significant differences in HRQOL loss across countries, after adjustment for diagnosis and sociodemographic factors.

**Conclusion:** HRQOL decreased with diagnosis severity, with significant differences between countries. To conduct cost-effectiveness modeling on HPV preventive interventions, obtaining accurate HRQOL estimates is essential. This process should consider the diverse local health preferences influenced by sociodemographic and cultural factors, as well as the population’s beliefs and experiences regarding health.

**PLAIN ENGLISH SUMMARY:** Human papillomavirus (HPV) infection can cause precancerous lesions and cervical cancer, but there is limited information about how these conditions affect women’s quality of life, especially in low- and middle-income countries where the disease burden is highest. This study aimed to measure quality of life among women with HPV-positive test results, precancerous cervical lesions, and cervical cancer in Latin America.

We included 1,073 women aged 18 to 75 years from Bolivia, Colombia, Honduras, Peru, and Uruguay. Participants completed the EQ-5D-5L, an internationally validated questionnaire used to measure health-related quality of life.

We found that quality of life worsened as disease severity progressed. Women with cervical cancer experienced a 3.19-fold greater loss in quality of life compared with women who had an HPV-positive test result or mild precancerous lesions (CIN1). We also found important differences between countries.

These findings provide locally relevant quality-of-life estimates that can be used in future economic evaluations to help identify the most efficient strategies for preventing and treating cervical cancer and precancerous lesions in Latin America.

## BACKGROUND

Human papillomavirus (HPV) infection is endemic worldwide, with an estimated global prevalence of 11.7% in women with normal cytology, and important variations across regions [1]. Bruni et al., estimated cervical HPV prevalence in Latin America and the Caribbean to be 16.1% (95% confidence interval [CI]: 15.8% to 16.4%) [2]. HPV infection was responsible for 4.3% of all new cancers worldwide in 2020 [3]; it is found in all cervical cancers, in 43% to 88% of other anogenital cancers, and in 13% to 56% of oropharyngeal cancers [4]. In 2021, there were 28.2 cervical cancer cases per 100,000 women (95% CI: 25.5 to 31.1); and 10.8 associated deaths per 100,000 women (95% CI: 9.8 to 11.8) in the region [5]. Moreover, cervical cancer is responsible for 1,093,576 disability-adjusted life years lost in Latin America and the Caribbean [5]. Several studies have measured health-related quality of life (HRQOL) in women with HPV positive results and associated diseases [6–15]. HPV-positive results can negatively affect HRQOL in emotional, social, and sexual functioning areas. Women with a positive HPV test result must not only deal with the stigma associated with a sexually transmitted infection but also undergo further examinations, which cause fear, distress, and embarrassment [11, 16]. Women with HPV-related disease report physical and emotional changes in their lives, as well as changes in their sexual and affective relationships, and in their social relationships [17, 18]. Women with cervical cancer exhibit lower HRQOL scores compared to the general population, with a further decline during treatment [7, 11, 12, 19, 20].

Despite growing evidence on HRQOL in HPV-related diseases, there are few studies from Latin America to our knowledge, and those available primarily focus on cervical cancer or specific subdomains of HRQOL, such as sexual function [14, 15, 17, 21]. Understanding the impact of HPV test results and HPV-related diseases on women’s HRQOL in this region is critical for two key reasons. First, it enables more comprehensive care planning by accounting not only for clinical outcomes but also for the broader effects of the disease and its treatment on patients’ daily lives. Second, utility measures are essential for health economic evaluations, helping decision-makers identify the most efficient strategies for preventing and managing HPV-related diseases. It has been well-documented that the impact of disease on HRQOL can vary significantly across countries. The population’s perception of health is influenced by differences in socioeconomic and cultural factors, and the healthcare system [21–23]. As a result, findings from studies conducted in other regions may not directly apply to Latin America. This underscores the need for region-specific HRQOL research to better inform local health policies and interventions.

We aimed to describe HRQOL in women with a positive HPV test result or HPV-related cervical disease, providing utilities that can be used for future cost-effectiveness analyses of public health and clinical interventions aimed at preventing HPV infections and controlling cervical cancer. We also conducted an exploratory analysis of differences in HRQOL across diagnoses and countries.

## METHODOLOGY

### Study design

Our study was conducted in five Latin American countries: Bolivia, Colombia, Honduras, Peru, and Uruguay. Beginning in January 2022, we enrolled women aged 18 to 75 years with a recent diagnosis (no more than six months since the diagnosis was communicated to the patient) of an HPV-positive test detected through cervical cancer screening, cervical intraepithelial neoplasia (CIN), or first-time cervical cancer. We defined HPV test positivity as a positive HPV DNA test result, either by Hybrid Capture II (Qiagen, USA) or by the COBAS 4800 HPV Test (Roche Diagnostics, USA). CIN and cervical cancers were diagnosed histologically by local pathologists based on biopsy specimens collected during the colposcopic evaluation. We included only recent diagnoses because it has been reported that the period directly after the diagnosis is when patients experience the most acute decline in their emotional functioning [11]. We excluded women receiving treatment for cervical cancer or who had a disease that might significantly affect their HRQOL, such as other cancers or rheumatological diseases. We identified women at clinical sites from laboratory registers or among those who participated or were participating in the ESTAMPA study [24]. The study had two objectives: 1) to measure the HRQOL of women with HPV-associated health outcomes using a generic (EQ-5D-5L) questionnaire; 2) to measure the psychosocial impact of receiving a positive HPV test result, as well as the HRQOL in women with cancer, using disease-specific questionnaires.

### Sample size

For the first objective, we estimated that a sample size of at least 42 women per diagnosis was sufficient for a 10% precision in the EQ-5D index scores, assuming a 0.3 population standard deviation [12], and therefore aimed to recruit at least 210 women per country and at least 1,050 women in total. The current analysis focuses on this first objective. Enrollment is still ongoing, as a larger sample of women with HPV positive tests and cervical cancer was calculated for the second study objective, which will be reported in a separate manuscript.

### Data collection

We collected data on sociodemographic variables using a structured questionnaire administered through face-to-face interviews and chart review, including age, educational level, parity, number of lifetime sexual partners, and whether they had a current sexual partner. Clinical questions included diagnosis and cancer stage according to the International Federation of Gynecology and Obstetrics (FIGO) 2018 classification [25]. Additionally, all participants completed the EQ-5D-5L questionnaire (EuroQol Group, Rotterdam, the Netherlands) [26], developed and adapted for each country, except in Honduras, where we used the version corresponding to Mexico, as a validated country-specific questionnaire was not available. Participants also completed the Psycho-ESTAMPA [27] and FACT-CX [28] questionnaires, which will be reported separately once enrolment for objective two is complete.

The EQ-5D-5L instrument has been extensively described and used [29–31]. Briefly, it asks participants to assess their health state on the day of the interview in five domains: mobility, self-care, usual activities, pain/discomfort, and anxiety/depression. Each domain has five levels on a Likert scale: 1: no problems; 2: mild; 3: moderate; 4: severe; and 5: extreme problems. There are 3,125 possible health states, derived from all possible combinations of levels across the domains, ranging from 11111 (no problems in any dimension) to 55555 (extreme problems in all dimensions). The health states have a corresponding value for each country, obtained using the EuroQol Valuation Technology questionnaire in a general reference population [32–34]. The questionnaire also includes a visual analogue scale (VAS), which assigns a value of 0 to the worst possible health and 100 to the best health the participant can imagine [31, 35].

Researchers at each site entered the questionnaires into the Research Electronic Data Capture (REDCap) platform, and quality controls were performed regularly. To calculate the EQ-5D index scores, we first measured women’s health states using the EQ-5D-5L questionnaire; then, we translated these health states into the corresponding value. For participants enrolled in Peru and Uruguay, we were able to use country-specific value sets [33, 34]. For Bolivia, Colombia and Honduras, we used a reference value set corresponding to the non-English Hispanic population in the United States of America [32], as these countries lack local value sets. We used this value set because a previous study has shown higher differences between South American countries than those from a culturally comparable population of combined origin [36].

### Statistical analysis

We calculated the EQ-5D index score and VAS score medians and interquartile ranges (IQRs) for each diagnosis (HPV-positive test, CIN1, CIN2, CIN3, and cervical cancer) in each country, and the mean and standard deviation (SD) for each diagnosis across all countries. We also calculated the proportion of women reporting any problem (e.g., values from 2 to 5) in each dimension, as well as in at least one of the five dimensions, by country and diagnosis. Moreover, we calculated the proportion of women reporting perfect health (i.e., an EQ-5D-5L health state of 11111) by country and diagnosis.

The EQ-5D index can range from minus infinity to 1 and usually has a left-skewed distribution. The assumption of normality of residuals in multiple linear regression was not met; thus, we transformed the EQ-5D index score to fit a gamma distribution. We subtracted each score from 1, yielding a distribution from 0 to infinity. This transformation expresses the EQ-5D scores in terms of their disutility (loss in perfect health), where 0 represents no disutility (perfect health), and increasing values represent higher disutility. With the new distribution, we fit a gamma regression model to explore if diagnosis and country of residence were predictors of HRQOL.

We fitted two gamma regression models: one adjusting for country and diagnosis (Model 1), and a second adjusting additionally for age, education, and having children (Model 2). We dichotomized age as less than 45 years and 45 years and older; the cut point of 45 years was selected as it was close to the mean age. We also fitted a model including an interaction term between diagnoses and country. We carried out a type III test ANOVA to explore if country and diagnosis were significant predictors in each model. We calculated the estimated marginal means (EMMs) using the *emmeans package* in R [37] and performed pairwise comparisons. To further assess the effect of diagnoses, we also fit models with diagnoses as an ordinal factor to assess linear trends. Finally, we conducted a logistic regression with a state of perfect health as the outcome and countries and diagnoses as predictors to estimate odds ratios (ORs) for perfect health with their respective 95% CIs.

Differences between countries are potentially attributable to differences in both the valuation of health states and the value sets used across countries. We aimed to assess whether the differences observed between countries could arise solely from differences in weights across the three valuation studies (Peru, Uruguay, and the non-English Hispanic population in the United States). To assess this, we selected the group of health states reported by participants from the country with the widest range of unique health states and calculated the EQ-5D index scores using the weights corresponding to the three available value sets. Similarly to our main analysis, we transformed the EQ-5D index scores to fit a gamma distribution and fitted a gamma mixed-effects model (package *glmmTMB* in R) [38] with a log link and a random intercept for health profile to account for repeated valuation of identical health profiles across different value sets. For all analyses, we considered a significance level of 0.05. We conducted the analysis in R version 4.3.3 [39]. Our study was reported following the STROBE guidelines [40].

### Ethical considerations

Our study was approved by the local institutional review boards in each country, the Research Ethics Office of the Faculty of Medicine and Health Sciences, McGill University, and the Ethics Committee of the International Agency for Research on Cancer (IARC). All participants signed an informed consent form before data collection. The data was collected anonymously. The analysis was conducted using a secure, anonymized, and password-protected database (REDCap) [41].

## RESULTS

We enrolled 1,075 women; two had incomplete EQ-5D-5L data and were therefore excluded from the analysis. Here, we present HRQOL results using the EQ-5D-5L from the remaining 1,073 women (Table 1). Participants had a median age of 43 years (IQR: 34-52), 65.9% had secondary or higher education, and 81.5% had children. Regarding the EQ-5D dimensions (Table 2), 70.4% of participants reported at least some problems in one of the five dimensions. Anxiety or depression was the most affected dimension, with 58.5% of participants reporting at least some problems, followed by pain or discomfort, with 42.0% of women reporting some pain or discomfort. The least affected dimension was self-care, where only 3.9% reported any problems. As expected, the proportion of women reporting at least some problems increased as the diagnosis worsened, from 64.2% in women with an HPV-positive test result alone or CIN1 to 85.1% in women with cervical cancer. Women living in Uruguay consistently reported more problems compared with women in other participating countries across all five dimensions.

**Table 1.** Participants’ demographic characteristics in five Latin American countries.

| Characteristics | Bolivia<br>(n=350) | Colombia<br>(n=238) | Honduras<br>(n=327) | Peru<br>(n=116) | Uruguay<br>(n=42) | Total<br>(n=1,073) |
| --- | --- | --- | --- | --- | --- | --- |
| Median age (interquartile range) | 42 (34 to 50) | 44 (37 to 54) | 46 (35 to 56) | 36 (30 to 45) | 42 (35 to 48) | 43 (34 to 52) |
| Education |  |  |  |  |  |  |
| None | 17 (4.9%) | 4 (1.7%) | 34 (10.4%) | 0 (0%) | 0 (0%) | 55 (5.1%) |
| Primary | 41 (11.7%) | 54 (22.7%) | 198 (60.6%) | 1 (0.9%) | 17 (40.5%) | 311 (29.0%) |
| Secondary or higher | 292 (83.4%) | 180 (75.6%) | 95 (29.1%) | 115 (99.1%) | 25 (59.5%) | 707 (65.9%) |
| Have children (>=1) | 290 (82.9%) | 202 (84.9%) | 303 (92.7%) | 42 (36.2%) | 38 (90.5%) | 875 (81.5%) |
| Diagnosis |  |  |  |  |  |  |
| HPV-positive test without disease<sup>a</sup> | 162 (46.3%) | 74 (31.1%) | 83 (25.4%) | 38 (32.8%) | 7 (16.7%) | 364 (33.9%) |
| CIN1 | 78 (22.3%) | 41 (17.2%) | 67 (20.5%) | 27 (23.3%) | 1 (2.4%) | 214 (19.9%) |
| CIN2 | 45 (12.9%) | 21 (8.8%) | 43 (13.1%) | 19 (16.4%) | 3 (7.1%) | 131 (12.2%) |
| CIN3 | 35 (10.0%) | 23 (9.7%) | 42 (12.8%) | 24 (20.7%) | 12 (28.6%) | 136 (12.7%) |
| Cervical cancer | 30 (8.6%) | 79 (33.2%) | 92 (28.1%) | 8 (6.9%) | 19 (45.2%) | 228 (21.2%) |

CIN: cervical intraepithelial neoplasia; HPV: human papillomavirus.
<sup>a</sup> We defined disease as cervical intraepithelial neoplasia or cervical cancer.

**Table 2.**
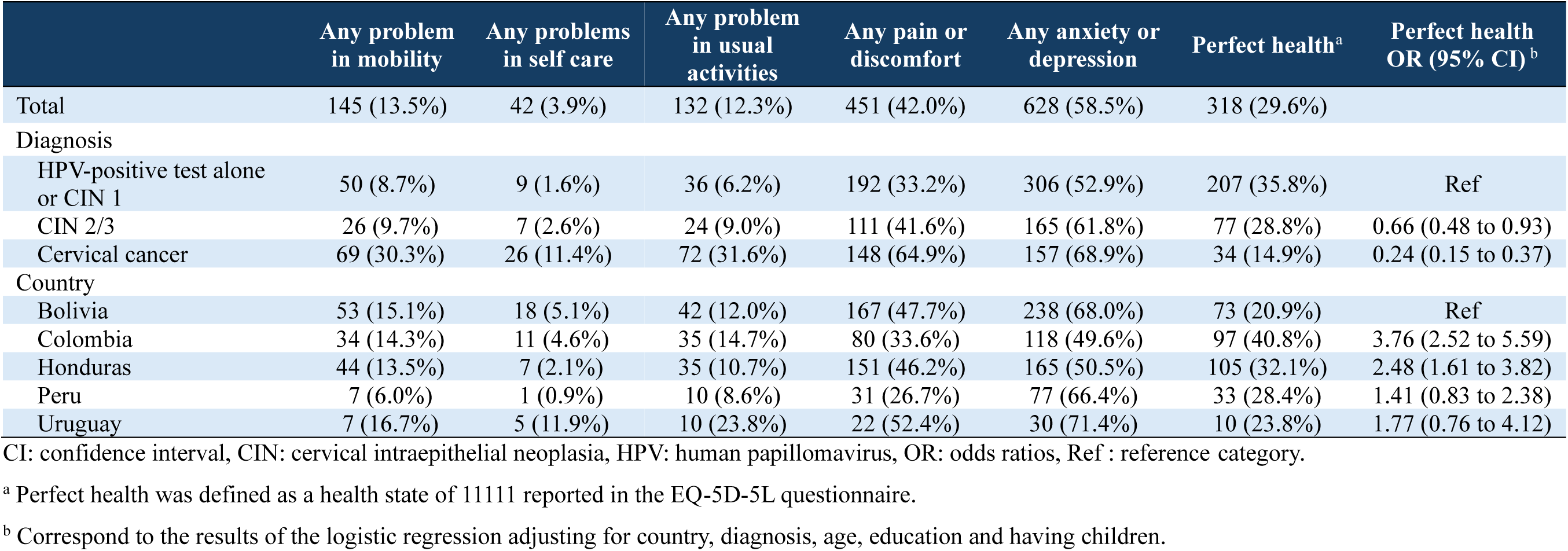
Presence of any health problems in five EQ domains and odds ratio for reporting perfect health, by country and diagnosis in women with an HPV-positive test and HPV-related disease from five Latin American countries.

|  | Any problem<br>in mobility | Any problems<br>in self care | Any problem<br>in usual<br>activities | Any pain or<br>discomfort | Any anxiety or<br>depression | Perfect health<sup>a</sup> | Perfect health<br>OR (95% CI)<sup>b</sup> |
| --- | --- | --- | --- | --- | --- | --- | --- |
| Total | 145 (13.5%) | 42 (3.9%) | 132 (12.3%) | 451 (42.0%) | 628 (58.5%) | 318 (29.6%) |  |
| Diagnosis |  |  |  |  |  |  |  |
| HPV-positive test alone<br>or CIN 1 | 50 (8.7%) | 9 (1.6%) | 36 (6.2%) | 192 (33.2%) | 306 (52.9%) | 207 (35.8%) | Ref |
| CIN 2/3 | 26 (9.7%) | 7 (2.6%) | 24 (9.0%) | 111 (41.6%) | 165 (61.8%) | 77 (28.8%) | 0.66 (0.48 to 0.93) |
| Cervical cancer | 69 (30.3%) | 26 (11.4%) | 72 (31.6%) | 148 (64.9%) | 157 (68.9%) | 34 (14.9%) | 0.24 (0.15 to 0.37) |
| Country |  |  |  |  |  |  |  |
| Bolivia | 53 (15.1%) | 18 (5.1%) | 42 (12.0%) | 167 (47.7%) | 238 (68.0%) | 73 (20.9%) | Ref |
| Colombia | 34 (14.3%) | 11 (4.6%) | 35 (14.7%) | 80 (33.6%) | 118 (49.6%) | 97 (40.8%) | 3.76 (2.52 to 5.59) |
| Honduras | 44 (13.5%) | 7 (2.1%) | 35 (10.7%) | 151 (46.2%) | 165 (50.5%) | 105 (32.1%) | 2.48 (1.61 to 3.82) |
| Peru | 7 (6.0%) | 1 (0.9%) | 10 (8.6%) | 31 (26.7%) | 77 (66.4%) | 33 (28.4%) | 1.41 (0.83 to 2.38) |
| Uruguay | 7 (16.7%) | 5 (11.9%) | 10 (23.8%) | 22 (52.4%) | 30 (71.4%) | 10 (23.8%) | 1.77 (0.76 to 4.12) |

CI: confidence interval, CIN: cervical intraepithelial neoplasia, HPV: human papillomavirus, OR: odds ratios, Ref : reference category.
<sup>a</sup> Perfect health was defined as a health state of 11111 reported in the EQ-5D-5L questionnaire.
<sup>b</sup> Correspond to the results of the logistic regression adjusting for country, diagnosis, age, education and having children.

A third (29.6%) of the respondents reported not having any problems in any of the five domains (e.g. perfect health defined as a health state 11111). In most countries, most women with perfect health were those with an HPV-positive test alone or a CIN1 diagnosis. However, some women with cervical cancer (14.9%) also reported not having problems in any of the five domains of the EQ-5D (Table 2). Women were less likely to report a state of perfect health if they had a diagnosis of CIN2/3 (OR: 0.66, 95% CI: 0.48 to 0.93) or cervical cancer (OR: 0.24, 95% CI: 0.15 to 0.37) compared to women with an HPV-positive test or CIN1. Adjusting for age, education, and having children did not change the magnitude of these associations. Women from Colombia (OR: 3.76, 95% CI: 2.52 to 5.59) and Honduras (OR: 2.48, 95% CI: 1.61 to 3.82) were more likely to report being in a state of perfect health than women from Bolivia. Similarly, women from Colombia (OR: 2.68, 95%CI: 1.53 to 4.68) were more likely to report being in a state of perfect health than women from Peru.

The mean EQ-5D index was 0.906 (SD: 0.129) for HPV-positive test alone, 0.891 (SD: 0.136) for CIN1, 0.892 (SD: 0.124) for CIN2, 0.870 (SD: 0.138) for CIN3, and 0.737 (SD: 0.268) for cervical cancer. There were no significant differences in EQ-5D index between HPV-positive test and CIN1, and between CIN2 and CIN3 (Supplement Table S1); therefore, we combined these diagnoses for analyses. We present the EQ-5D index (Figure 1) and the EQ-VAS (Supplement Figure S1) for each combined diagnosis and country in Table 3. In Supplement Table S2, we present EQ-5D index for diagnoses separately. We found a moderate correlation between EQ-5D index and the EQ-VAS (Spearman coefficient = 0.532, p<0.001). Type III ANOVA test showed that there was a significant effect of country of residence (p<0.001), diagnosis (p<0.001), age (p<0.001) and education (p<0.031), as well as interactions between country and diagnosis (p=0.013) on the EQ-5D index score; this indicates that there are both important differences between countries in reported HRQOL, and that the effect of diagnosis on HRQOL is different between countries.

**Fig. 1.**
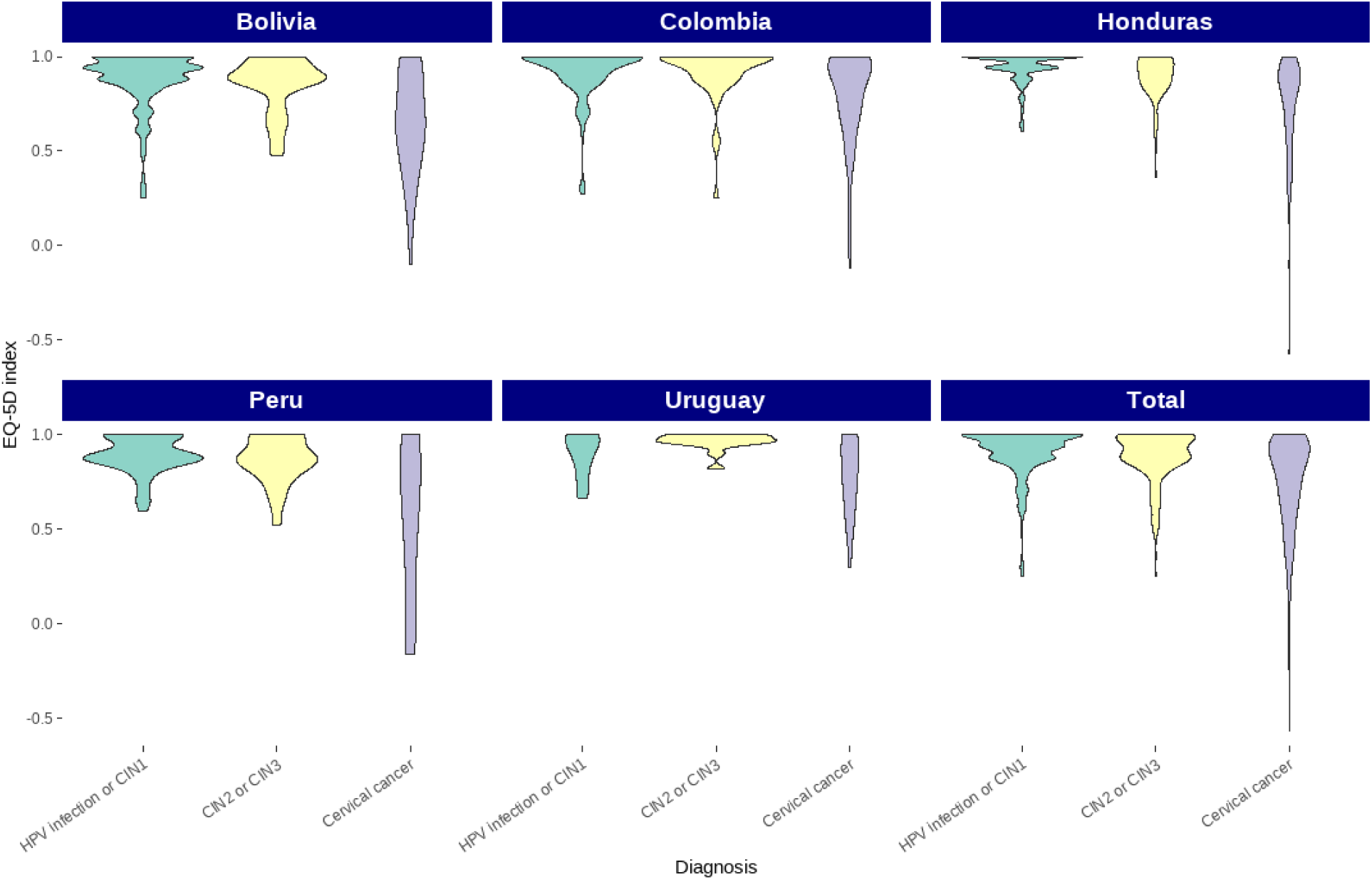
EQ-5D index distribution by country and diagnosis in five Latin American countries CIN: cervical intraepithelial neoplasia, HPV: Human Papillomavirus, EQ-5D: EuroQol 5 Dimension questionnaire.

**Table 3.** Health-related quality of life in women with an HPV-positive test and HPV-related disease in five Latin American countries.

|  | Bolivia<br>(n=350) | Colombia<br>(n=238) | Honduras<br>(n=327) | Peru<br>(n=116) | Uruguay<br>(n=42) | Total<br>(n=1,073) | HRQOL<br>loss ratio between<br>diagnoses (95%<br>CI)<sup>a</sup> |
| --- | --- | --- | --- | --- | --- | --- | --- |
| *Median (IQR) EQ-5D index score by diagnosis* |  |  |  |  |  |  |  |
| HPV-positive test alone or<br>CIN1 | 0.940<br>(0.845 to 0.943) | 0.943<br>(0.877 to 1.000) | 0.943<br>(0.940 to 1.000) | 0.877<br>(0.874 to 1.000) | 0.953<br>(0.840 to 1.000) | 0.943<br>(0.877 to 1.000) | Ref |
| CIN2/3 | 0.883<br>(0.807 to 0.943) | 1.000<br>(0.877 to 1.000) | 0.932<br>(0.845 to 1.000) | 0.877<br>(0.805 to 1.000) | 0.970<br>(0.961 to 1.000) | 0.883<br>(0.845 to 1.000) | 1.33 (1.11 to 1.59) |
| Cervical cancer | 0.641<br>(0.439 to 0.912) | 0.877<br>(0.682 to 0.972) | 0.845<br>(0.624 to 0.911) | 0.665<br>(0.319 to 0.890) | 0.777<br>(0.646 to 0.970) | 0.817<br>(0.610 to 0.943) | 3.19 (2.61 to 3.90) |
| *Median (IQR) EQ-VAS by diagnosis* |  |  |  |  |  |  |  |
| HPV-positive test alone or<br>CIN1 | 70 (60 to 80) | 90 (80 to 95) | 95 (80 to 100) | 80 (70 to 90) | 75 (58 to 83) | 80 (70 to 90) | NA |
| CIN2/3 | 70 (60 to 80) | 90 (80 to 95) | 85 (80 to 95) | 80 (67 to 88) | 90 (80 to 94) | 80 (70 to 90) | NA |
| Cervical cancer | 53 (40 to 80) | 80 (65 to 90) | 70 (55 to 80) | 70 (58 to 81) | 66 (50 to 85) | 70 (60 to 81) | NA |

CI: confidence interval CIN: cervical intraepithelial neoplasia, EQ-5D: EuroQol 5 Dimension questionnaire, EQ-VAS: EuroQol visual analogue scale, HPV: human papillomavirus, HRQOL: health-related quality of life, IQR: interquartile range, NA: not applicable, Ref: reference category.
<sup>a</sup> Correspond to the exponentiated coefficients from a gamma regression model adjusted for country, diagnosis, age, education, and having children.

The gamma regression model (Table 3, Supplemental Table S3) showed a decreasing trend in the EQ-5D index with worsening diagnosis in both models (p for a linear trend <0.001). Pairwise comparisons (Supplement Table S1) showed that, after adjusting for country, age, education and having children, women with cervical cancer had a 3.19-fold higher HRQOL loss (disutility) compared to women with an HPV-positive test alone or CIN1. Regarding differences between countries, we found greater HRQOL loss in Colombia (HRQOL loss ratio: 1.61; 95% CI: 1.21 to 2.15), Honduras (HRQOL loss ratio: 1.75; 95% CI: 1.30 to 2.35), and Uruguay (HRQOL loss ratio: 1.88; 95% CI: 1.07 to 3.32) compared to Bolivia, as well as in Colombia (HRQOL loss ratio: 1.54; 95% CI: 1.02 to 2.32), and Honduras (HRQOL loss ratio: 1.67; 95% CI: 1.09 to 2.56) compared to Peru.

In the sensitivity analysis, we used the 62 reported health states from Bolivia. We found significant differences in HRQOL loss between the three weight sets (Supplement Table S4). While the Peruvian value set estimated a greater loss (HRQOL loss ratio: 1.26; 95% CI: 1.16 to 1.37) in HRQOL than the Hispanic value set, the Uruguayan value set estimates a lower loss than the Hispanic (HRQOL loss ratio: 0.44; 95% CI: 0.41 to 0.48) and Peruvian value sets (HRQOL loss ratio: 0.35; 95% CI: 0.32 to 0.38).

## DISCUSSION

We found that HRQOL in women with an HPV-positive test and HPV-associated cervical diseases decreased with severity of diagnosis, with EQ-D index scores decreasing from 0.906 in women with an HPV-positive test alone to 0.737 in women with cervical cancer. Similarly, HRQOL was associated with age and country of residence, with Colombia, Honduras and Uruguay having higher HRQOL loss than Bolivia, and Colombia and Honduras having higher HRQOL loss than Peru.

Here, we present HRQOL measurements for HPV-positive test results and HPV-related diseases using the EQ-5D-5L instrument. This approach allows our results to be used in future health economic research assessing interventions for cervical cancer prevention in Latin America. This is especially relevant for cost-effectiveness studies being conducted in the context of the World Health Organization initiative for cervical cancer elimination [42]. To our knowledge, these are one of the first EQ-5D-5L-based HRQOL measurements for HPV-positive test results and HPV-related diseases conducted in these countries. Importantly, several health economics guidelines recommend the use of utilities derived from EQ-5D assessments, such as those we report [28–31].

In our study, the EQ-5D index decreased as the disease progressed. Previous studies have shown similar results [8, 10, 13, 21, 43, 44]. Galante et al., [21] reported that HRQOL in women with CIN and cervical cancer in Argentina and Chile decreases with the severity of the disease. de Kok et al., [13] reported EQ-5D index scores between 0.89 and 0.94 in the initial six months after referral to colposcopy in the Netherlands. An EQ-5D index score of 0.88 has been reported for low-grade abnormalities in studies conducted in other countries, [9] while the scores varied between 0.81 and 0.93 [12, 21] for CIN1, between 0.63 and 0.9 for CIN2/3 [12, 21]; and between 0.19 and 0.89 for cervical cancer [12, 13, 20, 21, 45, 46]. Similar results have also been observed using the SF-36 in Colombia [14].

Women in our study reported anxiety or depression as the most frequently affected domain in all diagnoses and in all countries. Women with HPV-positive screening test results may experience anxiety and concern regarding their condition; they may also feel anger, shame, and guilt, with a negative body image and a decline in self-esteem [11]. Nevertheless, as the disease progresses, women may start experiencing symptoms like pain in the case of cervical cancer, and the side effects of treatment, which further impact HRQOL. This is evident from studies conducted on women with cervical cancer in Ethiopia and India, where pain or discomfort was the most frequently affected EQ-5D domain, followed by difficulty in performing usual activities [45, 46]. HRQOL was significantly lower in women with CIN2/3 precancerous lesions compared to those with an HPV-positive test result or CIN1. This may be attributed to increased anxiety/depression due to the progression of the disease.

EQ-5D index scores are derived from two components: the health profile reported by the respondent and the corresponding value assigned to that profile using a country-specific value set derived from population-based preference studies. Therefore, differences between countries may arise not only from variations in how women perceive and report their current health, but also from differences in how populations value the same health states. Despite using the same value set across three countries (Bolivia, Honduras, and Colombia), we still observed differences among them. This suggests that part of the difference is attributable to differences in women’s perceptions of how their diagnosis has affected their health.

Socioeconomic and cultural factors, as well as factors related to the health system, affect how individuals perceive their health [9]. It has been reported that HRQOL, and more specifically, the levels of anxiety, are related to the way the health practitioner delivers the diagnosis [27]. Similarly, women with a better understanding of their health condition could show less anxiety and depression [47, 48]. We also found in our sensitivity analysis that the observed difference in HRQOL loss between countries could also be attributable to the value sets, similar to a previous report [21]. Country differences in value sets could be explained both through methodological variations in how the sets were derived (e.g., modeling strategy, translation of the questionnaire, quality control, and any imbalance in the sample composition) and inherent differences in health state preferences of the populations. Country-specific characteristics, such as healthcare systems, wealth, cultural norms, and even climate and geography, could also affect populations’ health preferences [23]. There could also be secular trends in population health preferences, given the important time gap between the valuation studies (for example, 15 years between Peru and US-Hispanic) [32–34].

An important consideration is whether the observed differences are clinically meaningful. Hu et al., [49] proposed a minimal clinically significant difference for an EQ-5D index of 0.039 (95% CI: 0.023 to 0.064) pre- and post-treatment. Pickard et al., [50] reported higher minimal important difference in patients with cancer as disease progresses in the United Kingdom and United States; for example, patients in the United Kingdom with advanced cancer stages had a minimal important difference of 0.16 compared to earlier stages. We found differences between diagnoses and countries that exceed these reference values, suggesting they are clinically significant.

The variability among different populations is a significant source of uncertainty in health economic evaluations, underscoring the importance of using local values. Kok et al. reported significant variations in utility scores for various health states related to cervical cancer screening [13]. They later compared these utility sets in a single micro-simulation and reached different conclusions regarding the most cost-effective screening test. What might be individually a small HRQOL loss could represent a substantial loss at the population level. Variations in utilities can significantly impact cost-effectiveness analysis conclusions, highlighting the need for locally derived values [14]. Moreover, it is important to include utilities for all relevant health states in a cost-effectiveness model; therefore, we collected data on HPV-positive test results and distinct precancerous lesions separately, in addition to cervical cancer.

We should acknowledge some limitations in our study. First, while we collected and adjusted analyses for some demographic and socioeconomic indicators, unmeasured variables may influence HRQOL, which we were unable to adjust for. Factors such as age, education, smoking, ethnicity and household income have been previously associated with HRQOL in women with HPV-related diseases [9, 19, 45]. We did not find an association between education and EQ-5D index scores. While we did adjust for age, education, and parity, we did not plan our sample size to examine these associations, nor did we collect data on household income or other variables that could affect HRQOL. Second, as we used convenience sampling, the women in our study might not represent the country’s health preferences. This limitation could be particularly relevant in Peru, where all women were enrolled in a private clinic; nevertheless, in the other countries, women were enrolled in public clinics receiving a wider range of patients. Thirdly, our study was designed to provide precise estimates of EQ-5D scores for each diagnosis and for each country separately. However, it was not powered to provide precise country-specific estimates within each diagnosis category. So, while the study is powered to assess aggregate differences between countries, differences within specific diagnostic categories should be interpreted with caution. Finally, we used a culturally comparable population of combined origin for those countries without a local value set; therefore, the calculated EQ-5D index scores may not fully represent these countries’ valuation. Nevertheless, we consider that using this combined value set would provide less biased results than using the value set from another country, as previous research has shown that a country’s valuations are likely to be closer to the Hispanic value set than to another country’s due to the known large differences which exist between countries [36].

Here, we found that HRQOL associated with HPV-positive test results and HPV-related diseases decreases as the disease progresses, with values going from 0.906 for an HPV-positive test without apparent disease to 0.737 for cervical cancer. We also found that HRQOL associated with an HPV-positive test and HPV-related cervical diseases varies markedly between countries. Evaluation of HRQOL is important for facilitating clinical decision-making to improve patient outcomes and for conducting health economic evaluations. Hence, the value of our study is that it provides researchers with local utility measures for the most relevant health states to use when conducting cost-effectiveness studies in Latin America, helping public health stakeholders decide on new interventions to prevent cervical cancer.

## HPV AND QUALITY OF LIFE IN LATIN AMERICA AND THE CARIBBEAN (HPVQOL-LAC) GROUP

Jafet Ortiz-Quintero (Instituto de Investigaciones en Microbiología, Universidad Nacional Autónoma de Honduras, Tegucigalpa, Honduras), Bettsy Flores and Osmalia Zambrana (Facultad de Medicina, Universidad Mayor, Real y Pontificia de San Francisco Xavier de Chuquisaca, Sucre, Bolivia), Laura Gomez (Fundación Santa Fé de Bogotá. Bogotá, Colombia), Natalia Perez (Centro Hospitalario Pereyra Rossell. Unidad Docente Clínica Ginecotocológica C, Montevideo, Uruguay), Valentina Florencia Bafico Rocha (Centro Hospitalario Pereyra Rossell. Unidad Docente Clínica Ginecotocológica C, Montevideo, Uruguay) and Sandra Liliana Martínez (Instituto Nacional de Cancerología. Bogotá, Colombia)

## AUTHORS CONTRIBUTION

RT and ATR conceptualized the study and developed the questionnaire. RT conducted the formal analysis and visualization. AF, YC, CT, VM, LT, YS, LG, JR, GV, MPV, GR, AB, and the other members of the HPVQoL-LAC Group were on charge of obtaining local ethical approval, enrolling participants, and collecting data. Analyses were validated by ATR, MA and TM. TM, MA and ELF supervised the work. RT prepared the original draft of the manuscript, and all authors contributed to interpretation of the results, the writing, review, and editing. All authors had access to the underlying data, verified the results, and approved the final version of the manuscript.

## DATA AVAILABILITY STATEMENT

The data used in this study selected several variables (age at recruitment, education, having children, screening history, cytology and HPV testing results, colposcopy, histology) from the “Quality of life in women recently diagnosed with HPV infection, precancerous cervical lesions and cervical cancer in Latin American countries study” database, which is stored securely at the International Agency for Research on Cancer. Anonymized individual participant data or aggregated data are available upon reasonable request to the corresponding author, after signing a contract, and with approval from the principal and local investigators.

## DISCLAIMER

MA is a staff member of the World Health Organization. The author alone is responsible for the views expressed in this article and she does not necessarily represent the decisions, policy or views of the World Health Organization. Where authors are identified as personnel of the International Agency for Research on Cancer/World Health Organization, the authors alone are responsible for the views expressed in this article and they do not necessarily represent the decisions, policy or views of the International Agency for Research on Cancer/World Health Organization. AB was supported by an appointment to the National Cancer Institute (NCI) Research Participation Program administered by the Oak Ridge Institute for Science and Education (ORISE) through an interagency agreement between the U.S. Department of Energy (DOE) and the National Institute of Health. ORISE is managed by ORAU under DOE contract number DESC0014664. All opinions expressed in this paper are the author’s and do not necessarily reflect the policies and views of NIH, NCI, DOE, or ORAU/ORISE. This research was supported in part by the Intramural Research Program of the National Institutes of Health (NIH). The contributions of the NIH author(s) are considered Works of the United States Government. The findings and conclusions presented in this paper are those of the author(s) and do not necessarily reflect the views of the NIH or the U.S. Department of Health and Human Services. Through his institution, ELF has received unconditional grants from Merck to complement funding received for his investigator-initiated studies on the epidemiology of HPV infection and cervical cancer. He has also served as an occasional advisor to Merck on HPV vaccination.

## Data Availability

The data used in this study selected several variables (age at recruitment, education, having children, screening history, cytology and HPV testing results, colposcopy, histology) from the “Quality of life in women recently diagnosed with HPV infection, precancerous cervical lesions and cervical cancer in Latin American countries” study database, which is stored securely at the International Agency for Research on Cancer. Anonymized individual participant data or aggregated data are available upon reasonable request to the corresponding author, after signing a contract, and with approval from the principal and local investigators.

## REFERENCES

1. Serrano, B., Brotons, M., Bosch, F.X., and Bruni, L. (2018). Epidemiology and burden of HPV-related disease. Best Pract Res Clin Obstet Gynaecol, 47,14–26.

2. Bruni, L., Diaz, M., Castellsague, X., Ferrer, E., Bosch, F.X., and de Sanjose, S. (2010). Cervical human papillomavirus prevalence in 5 continents: meta-analysis of 1 million women with normal cytological findings. J Infect Dis, 202(12), 1789–1799.

3. 3. International Agency for Research on Cancer, World Health Organization (2020). Cancers atributable to infections. Retrieved March 18, 2025, from https://gco.iarc.fr/causes/infections/tools-pie?mode=2&sex=0&population=who&continent=0&country=0&population_group=0&cancer=0&key=attr_cases&lock_scale=0&pie_mode=1&nb_results=5.

4. 4. de Martel, C., Ferlay, J., Franceschi, S., Vignat, J., Bray, F., Forman, D., et al. (2012). Global burden of cancers attributable to infections in 2008: a review and synthetic analysis. Lancet Oncol, 13(6), 607–615.

5. 5. Global Burden of Disease Collaborative Network. (2017) Global Burden of Disease Study. Results. Retrived March 18, 2025, from https://ghdx.healthdata.org/gbd-2017

6. 6. Cianci, S., Tarascio, M., Arcieri, M., La Verde, M., Martinelli, C., Capozzi, V.A., et al. (2023). Post Treatment Sexual Function and Quality of Life of Patients Affected by Cervical Cancer: A Systematic Review. Medicina (Kaunas), 59(4), 704.

7. Pfaendler, K.S., Wenzel, L., Mechanic, M.B., and Penner, K.R. (2015). Cervical cancer survivorship: long-term quality of life and social support. Clin Ther, 37(1), 39–48.

8. Zhao, M., Pu, X., Yan, Y.J., Zhang, S., Long, X., Luo, L., et al. (2021). The quality of life in women with cervical cancer and precancerous lesions of Han and ethnic minorities in Southwest China. BMC Cancer, 21(1), 1110.

9. Whynes, D.K. and TOMBOLA Group. (2008). Correspondence between EQ-5D health state classifications and EQ VAS scores. Health Qual Life Outcomes, 6(94).

10. Somanna, S.N., Sastry, N.B., Chaluvarayaswamy, R., and Malila, N. (2022). Quality of life and Its Determinants among Cervical Cancer Patients in South India. Asian Pac J Cancer Prev, 23(8), 2727–2733.

11. Fleurence, R.L., Dixon, J.M., Milanova, T.F., and Beusterien, K.M. (2007). Review of the economic and quality-of-life burden of cervical human papillomavirus disease. Am J Obstet Gynecol, 196(3), 206–212.

12. Marcellusi, A., Capone, A., Favato, G., Mennini, F.S., Baio, G., Haeussler, K., et al. (2015). Health utilities lost and risk factors associated with HPV-induced diseases in men and women: the HPV Italian collaborative study group. Clin Ther, 37(1), 156–167.

13. 13. de Kok, I., Korfage, I.J., van den Hout, W.B., Helmerhorst, T.J.M., Habbema, J.D.F., Essink-Bot, M.L., et al. (2018). Quality of life assumptions determine which cervical cancer screening strategies are cost-effective. Int J Cancer, 142(11), 2383–2393.

14. Urrea Cosme, Y., Cordoba Sanchez, V., Sanchez, G.I., Baena, A., Ruiz Osorio, M.A., Rodriguez Zabala, D., et al. (2020). Health-related quality of life of women after HPV testing as triage strategy for an abnormal Pap smear: a nested randomized pragmatic trial in a middle-income country. Qual Life Res, 29(11), 2999–3008.

15. Garces-Palacio, I.C., Sanchez, G.I., Baena Zapata, A., Cordoba Sanchez, V., Urrea Cosme, Y., Rodriguez Zabala, D., et al. (2020). Psychosocial impact of inclusion of HPV test on the management of women with atypical squamous cells of undetermined significance: a study within a randomised pragmatic trial in a middle-income country. Psychol Health, 35(6), 750–769.

16. Coronado, P.J., Gonzalez-Granados, C., Ramirez-Mena, M., Calvo, J., Fasero, M., Bellon, M., et al. (2022). Development and psychometric properties of the human papillomavirus-quality of life (HPV-QoL) questionnaire to assess the impact of HPV on women health-related-quality-of-life. Arch Gynecol Obstet, 306(4), 1085–1100.

17. Pereira-Caldeira, N.M.V., Goes, F.G.B., Almeida-Cruz, M.C.M., Caliari, J.S., Pereira-Avila, F.M.V., and Gir, E. (2020). Quality of Life for Women with Human Papillomavirus-induced Lesions. Rev Bras Ginecol Obstet, 42(4), 211–217.

18. Wang, S.M., Shi, J.F., Kang, D.J., Song, P., Qiao, Y.L., and Chinese, H.P.V.S.G. (2011). Impact of human papillomavirus-related lesions on quality of life: a multicenter hospital-based study of women in Mainland China. Int J Gynecol Cancer, 21(1), 182–188.

19. Osann, K., Hsieh, S., Nelson, E.L., Monk, B.J., Chase, D., Cella, D., et al. (2014). Factors associated with poor quality of life among cervical cancer survivors: implications for clinical care and clinical trials. Gynecol Oncol, 135(2), 266–272.

20. Lang, H.C., Chuang, L., Shun, S.C., Hsieh, C.L., and Lan, C.F. (2010). Validation of EQ-5D in patients with cervical cancer in Taiwan. Support Care Cancer, 18(10), 1279–1286.

21. Galante, J., Augustovski, F., Colantonio, L., Bardach, A., Caporale, J., Marti, S.G., et al. (2011). Estimation and comparison of EQ-5D health states’ utility weights for pneumococcal and human papillomavirus diseases in Argentina, Chile, and the United Kingdom. Value Health, 14(5 Suppl 1), S60–64.

22. Konig, H.H., Bernert, S., Angermeyer, M.C., Matschinger, H., Martinez, M., Vilagut, G., et al. (2009). Comparison of population health status in six european countries: results of a representative survey using the EQ-5D questionnaire. Med Care, 47(2), 255–261.

23. Roudijk, B.; Janssen, B.; Olsen, J.A. (2022). How Do EQ-5D-5L Value Sets Differ? In N. Devlin, B. Roudijk, & K. Ludwig (Eds.), Value Sets for EQ-5D-5L. A Compendium, Comparative Review & User Guide (pp. 235–258). Springer, Cham.

24. Almonte, M., Murillo, R., Sánchez, G.I., González, P., Ferrera, A., Picconi, M.A., et al. (2020). Multicentric study of cervical cancer screening with human papillomavirus testing and assessment of triage methods in Latin America: the ESTAMPA screening study protocol. BMJ open, 10(5), e035796.

25. Bhatla, N., Aoki, D., Sharma, D.N., and Sankaranarayanan, R. (2018). Cancer of the cervix uteri. Int J Gynaecol Obstet, 143(Suppl 2), 22–36.

26. 26. EuroQol Research Foundation (2021). *EQ-5D-5L*. EuroQol Instruments. Retrieved July 11, 2024, from: https://euroqol.org/information-and-support/euroqol-instruments/eq-5d-5l/.

27. Arrossi, S., Almonte, M., Herrero, R., Gago, J., Sanchez Antelo, V., Szwarc, L., et al. (2020). Psycho-social impact of positive human papillomavirus testing in Jujuy, Argentina results from the Psycho-Estampa study. Prev Med Rep, 18, 101070.

28. Peerawong, T., Suphasynth, Y., Kongkamol, C., Rordlamool, P., Bridhikitti, J., Jiratrachu, R., et al. (2020). Validation of the Functional Assessment of Cancer Therapy with Cervical Cancer Subscale (FACT-CX) for Quality of Life in Thai Patients Prior to Chemoradiotherapy. Asian Pac J Cancer Prev, 21(7), 1891–1897.

29. Feng, Y.S., Kohlmann, T., Janssen, M.F., and Buchholz, I. (2021). Psychometric properties of the EQ-5D-5L: a systematic review of the literature. Qual Life Res, 30(3), 647–673.

30. 30. Rabin, R. and de Charro, F. (2001). EQ-5D: a measure of health status from the EuroQol Group. Ann Med, 33(5), 337–343.

31. Roudijk, B., Ludwig, K., and Devlin, N. (2022). EQ-5D-5L Value Set Summaries. In N. Devlin, B. Roudijk, & K. Ludwig (Eds.), Value Sets for EQ-5D-5L. A Compendium, Comparative Review & User Guide, (pp. 55–212). Springer, Cham.

32. Zarate, V., Kind, P., and Chuang, L.-H. (2008). Hispanic Valuation of the EQ-5D Health States: A Social Value Set for Latin Americans. Value in Health, 11(7), 1170–1177.

33. Augustovski, F., Belizán, M., Gibbons, L., Reyes, N., Stolk, E., Craig, B., et al. (2020). Peruvian Valuation of the EQ-5D-5L: a direct comparison of TTO and DCE. Value Health, 23(7), 880–888.

34. Augustovski, F., Rey-Ares, L., Irazola, V., Garay, O.U., Gianneo, O., Fernandez, G., et al. (2016). An EQ-5D-5L value set based on Uruguayan population preferences. Qual Life Res, 25(2), 323–333.

35. 35. EuroQol Research Foundation (2019). EQ-5D-5L User Guide. Retrieved March 18, 2025, from https://euroqol.org/publications/user-guides.

36. Tejada, R.A., Gibbons, L., Belizan, M., Gutierrez, E.L., Reyes, N., and Augustovski, F.A. (2021). Comparison of EQ-5D Values Sets Among South American Countries. Value Health Reg Issues, 26, 56–65.

37. 37. Lenth, R. (2024). emmeans: Estimated Marginal Means, aka Least-Squares Means. R package. Retrieved March 18, 2025, from https://github.com/rvlenth/emmeans/

38. 38. Brooks, M.E., Kristensen, K., van Benthem, K. J., Magnusson, A., Berg, C. W., Nielsen, A., Skaug, H. J., Mächler, M., & Bolker, B. M. (2017). glmmTMB balances speed and flexibility among packages for zero-inflated generalized linear mixed modeling. The R Journal, 9(2), 378–400.

39. R: The R Project for Statistical Computing. Retrieved March 18, 2025, from https://www.r-project.org/

40. von Elm, E., Altman, D.G., Egger, M., Pocock, S.J., Gotzsche, P.C., Vandenbroucke, J.P., et al. (2007). The Strengthening the Reporting of Observational Studies in Epidemiology (STROBE) statement: guidelines for reporting observational studies. Ann Intern Med, 147(8), 573–577.

41. Harris, P.A., Taylor, R., Minor, B.L., Elliott, V., Fernandez, M., O’Neal, L., et al. (2019). The REDCap consortium: Building an international community of software platform partners. J Biomed Inform, 95, 103208.

42. 42. World Health Organization. (2020). Global strategy to accelerate the elimination of cervical cancer as a public health problem. Geneva, Switzerland. p. 56. Retrieved March 18, 2025, from https://www.who.int/publications/i/item/9789240014107

43. Ju, X., Canfell, K., Howard, K., Garvey, G., Hedges, J., Smith, M., et al. (2021). Population-based utility scores for HPV infection and cervical squamous cell carcinoma among Australian Indigenous women. PLoS One, 16(7), e0254575.

44. Hung, M.C., Wu, C.L., Hsu, Y.Y., Hwang, J.S., Cheng, Y.M., and Wang, J.D. (2014). Estimation of potential gain in quality of life from early detection of cervical cancer. Value Health, 17(4), 482–486.

45. Jyani, G., Chauhan, A.S., Rai, B., Ghoshal, S., Srinivasan, R., and Prinja, S. (2020). Health-related quality of life among cervical cancer patients in India. Int J Gynecol Cancer, 30(12), 1887–1892.

46. Araya, L.T., Fenta, T.G., Sander, B., Gebremariam, G.T., and Gebretekle, G.B. (2020). Health-related quality of life and associated factors among cervical cancer patients at Tikur Anbessa specialized hospital, Addis Ababa, Ethiopia. Health Qual Life Outcomes, 18(1), 72.

47. Sharp, L.K., Zurawski, J.M., Roland, P.Y., O’Toole, C., and Hines, J. (2002). Health literacy, cervical cancer risk factors, and distress in low-income African-American women seeking colposcopy. Ethn Dis, 12(4), 541–6.

48. Lee Mortensen, G. and Adeler, A.L. (2010). Qualitative study of women’s anxiety and information needs after a diagnosis of cervical dysplasia. Z Gesundh Wiss, 18(5), 473–482.

49. Hu, X., Jing, M., Zhang, M., Yang, P., and Yan, X. (2020). Responsiveness and minimal clinically important difference of the EQ-5D-5L in cervical intraepithelial neoplasia: a longitudinal study. Health Qual Life Outcomes, 18(1), 324.

50. Pickard, A.S., Neary, M.P., and Cella, D. (2007). Estimation of minimally important differences in EQ-5D utility and VAS scores in cancer. Health Qual Life Outcomes, 5(70).

