## Supplemental Information for "Quality of life in women with a human papillomavirus-positive screening test, cervical neoplasia, or cervical cancer in Latin America"

### Supplementary information

Romina A. Tejada, Arianis Tatiana Ramirez, Annabelle Ferrera, Yessy Cabrera, Carolina Terán, Vania Murillo, Lina Trujillo, Yuli Salgado, Juliana Rodriguez, Mateo Barros, Gino Venegas, María del Pilar Vera, Armando Baena, Guillermo Rodriguez, Andrea Beracochea, Eduardo L. Franco, Maribel Almonte, Talía Malagón, on behalf of the HPVQoL-LAC Group

**Corresponding author:**
Romina A. Tejada

Gerald Bronfman Department of Oncology, McGill University, Montreal, Canada

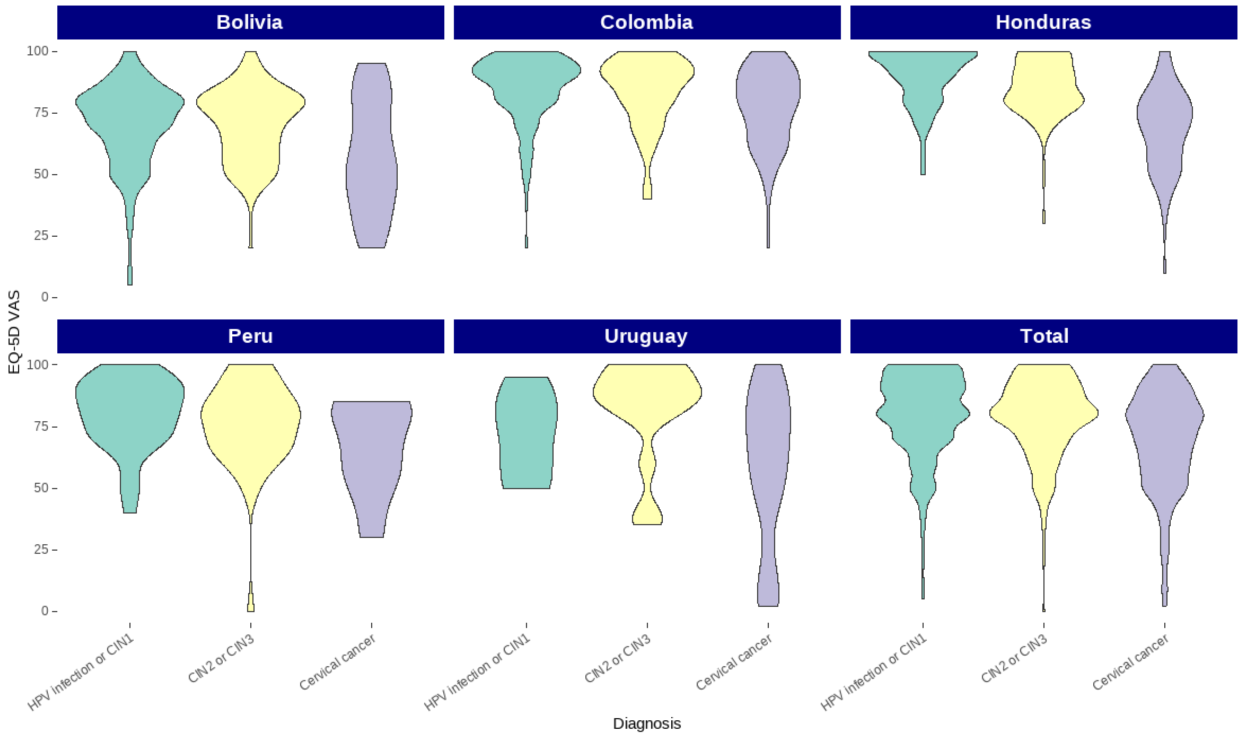


**Fig. S1** Visual analogue scale score distribution by country and diagnosis in five Latin American countries

CIN: cervical intraepithelial neoplasia, EQ-5D VAS: EuroQol visual analogue scale, HPV: human papillomavirus.

**Table S1.** Pairwise comparison in quality-of-life losses between diagnosis and between countries of residence using the EQ-5D index score.

| | Quality of life loss ratio (95% CI) from Gamma Model 1[a] | Quality of life loss ratio (95% CI) from Gamma Model 2[b] |
|---|---|---|
| ***Diagnosis*** | | |
| HPV-positive test alone to CIN1 | 1.27 (0.95 to 1.69) | 1.31 (0.98 to 1.75) |
| HPV-positive test alone to CIN2 | 1.24 (0.88 to 1.74) | 1.32 (0.93 to 1.86) |
| HPV-positive test alone to CIN3 | 1.58 (1.13 to 2.23) | 1.67 (1.19 to 2.37) |
| HPV-positive test alone to cervical cancer | 3.71 (2.77 to 4.99) | 3.60 (2.66 to 4.87) |
| CIN1 to CIN2 | 0.97 (0.67 to 1.41) | 1.01 (0.69 to 1.46) |
| CIN1 to CIN3 | 1.25 (0.86 to 1.81) | 1.28 (0.88 to 1.86) |
| CIN1 to cervical cancer | 2.93 (2.11 to 4.07) | 2.75 (1.97 to 3.84) |
| CIN2 to CIN3 | 1.28 (0.85 to 1.93) | 1.27 (0.84 to 1.92) |
| CIN2 to cervical cancer | 3.01 (2.07 to 4.37) | 2.73 (1.87 to 3.99) |
| CIN3 to cervical cancer | 2.35 (1.62 to 3.39) | 2.15 (1.48 to 3.12) |
| ***Combined diagnosis*** | | |
| HPV-positive test alone or CIN1 to CIN2/3 | 1.28 (1.03 to 1.58) | 1.33 (1.07 to 1.65) |
| HPV-positive test alone or CIN1 to cervical cancer | 3.35 (2.64 to 4.23) | 3.19 (2.50 to 4.06) |
| CIN2/3 to cervical cancer | 2.62 (2.01 to 3.42) | 2.40 (1.83 to 3.14) |
| ***Countries*** | | |
| Bolivia - Colombia | 1.67 (1.25 to 2.22) | 1.61 (1.21 to 2.15) |
| Bolivia - Honduras | 1.82 (1.40 to 2.37) | 1.75 (1.30 to 2.35) |
| Bolivia - Peru | 1.14 (0.80 to 1.64) | 1.05 (0.71 to 1.54) |
| Bolivia - Uruguay | 2.11 (1.21 to 3.69) | 1.88 (1.07 to 3.32) |
| Colombia - Honduras | 1.09 (0.82 to 1.45) | 1.09 (0.79 to 1.48) |
| Colombia - Uruguay | 1.27 (0.72 to 2.22) | 1.17 (0.66 to 2.07) |
| Honduras - Uruguay | 1.16 (0.67 to 2.01) | 1.08 (0.61 to 1.89) |
| Peru-Colombia | 1.46 (0.99 to 2.15) | 1.54 (1.02 to 2.32) |
| Peru-Honduras | 1.60 (1.11 to 2.30) | 1.67 (1.09 to 2.56) |
| Peru-Uruguay | 1.85 (1.00 to 3.40) | 1.80 (0.95 to 3.40) |

CI: confidence interval, HPV: human papillomavirus, CIN: cervical intraepithelial neoplasia.

[a] Model 1 adjusted by country and diagnosis.

[b] Model 2 adjusted by country, diagnosis, age, education and having children

**Table S2.** Health-related quality of life in women with an HPV-positive test and HPV-related disease in five Latin American countries by diagnosis.

| | Bolivia (n=350) | Colombia (n=238) | Honduras (n=327) | Peru (n=116) | Uruguay (n=42) | Total (n=1,073) |
|---|---|---|---|---|---|---|
| HPV-positive test alone | 0.940 (0.846 to 0.943) | 1.000 (0.888 to 1.000) | 1.000 (0.942 to 1.000) | 0.877 (0.874 to 1.000) | 0.978 (0.846 to 1.000) | 0.943 (0.877 to 1.000) |
| CIN1 | 0.883 (0.817 to 0.943) | 0.943 (0.845 to 1.000) | 0.943 (0.940 to 1.000) | 0.877 (0.877 to 1.000) | 0.866 [a] | 0.943 (0.877 to 1.000) |
| CIN2 | 0.883 (0.845 to 0.940) | 1.000 (0.877 to 1.000) | 0.943 (0.883 to 1.000) | 0.877 (0.843 to 1.000) | 0.970 (0.896 to 0.974) | 0.928 (0.874 to 1.000) |
| CIN3 | 0.877 (0.734 to 0.943) | 1.000 (0.877 to 1.000) | 0.883 (0.817 to 0.943) | 0.874 (0.802 to 0.908) | 0.970 (0.961 to 1.000) | 0.883 (0.817 to 1.000) |
| Cervical cancer | 0.641 (0.439 to 0.912) | 0.877 (0.682 to 0.972) | 0.845 (0.624 to 0.911) | 0.665 (0.319 to 0.890) | 0.777 (0.646 to 0.970) | 0.817 (0.610 to 0.943) |

HPV: human papillomavirus, CIN: cervical intraepithelial neoplasia.

[a] There was only one participant in this category.

Note: Values correspond to median (interquartile range).

**Table S3.** Factors associated to EQ-5D index in the gamma regression model.

| | Quality of life loss ratio[a] | 95% Confidence interval | P value |
|---|---|---|---|
| Bolivia | Ref | | |
| Colombia | 0.62 | 0.50 to 0.76 | **<0.001** |
| Honduras | 0.57 | 0.46 to 0.71 | **<0.001** |
| Peru | 0.95 | 0.72 to 1.26 | 0.741 |
| Uruguay | 0.53 | 0.35 to 0.80 | **0.002** |
| HPV-positive test or CIN1 | Ref | | |
| CIN2/3 | 1.33 | 1.11 to 1.59 | **0.002** |
| Cervical cancer | 3.19 | 2.61 to 3.90 | **<0.001** |
| Age 44 or younger | Ref | | |
| Age 45 or older | 1.46 | 1.24 to 1.72 | **<0.001** |
| No education | Ref | | |
| Primary | 0.70 | 0.49 to 1.01 | 0.054 |
| Secondary or higher | 0.89 | 0.62 to 1.28 | 0.518 |
| Not having children | Ref | | |
| Having children | 1.07 | 0.85 to 1.33 | 0.565 |

Ref: reference category.

[a] Correspond to the exponentiated coefficients from the gamma model adjusting for country, diagnosis, age, education and having children.

**Table S4.** HRQOL pairwise comparison between different weights sets.

| Value sets comparison | HRQOL loss ratio (95% CI) [a] | P value |
|---|---|---|
| Hispanic - Peru | 1.26 (1.16 to 1.37) | <0.001 |
| Hispanic - Uruguay | 0.44 (0.41 to 0.48) | <0.001 |
| Peru - Uruguay | 0.35 (0.32 to 0.38) | <0.001 |

HRQOL: health related quality of life; CI: confidence interval.

[a] Values were obtained using gamma mixed-effects model with a log link and a random intercept for health profile to account for repeated valuation of identical health states.
